# Assessment of Regional Body Composition, Physical Function and Sarcopenia among Peruvian women aging with HIV

**DOI:** 10.1101/2022.06.28.22277030

**Authors:** Diego M. Cabrera, Mijahil P. Cornejo, Yvett Pinedo, Patricia J. Garcia, Evelyn Hsieh

## Abstract

**Background:** Management of chronic conditions and optimization of overall health has become a primary global health concern in the care of people living with HIV, particularly in lower-and-middle income countries where infrastructure for chronic disease management may be fragmented. Alterations in body composition (BC) can reflect important changes in musculoskeletal health, particularly among populations at risk for developing fat and muscle redistribution syndromes, such as HIV-infected women. We aimed to explore this issue among Peruvian women aging with HIV.

**Methods:** Cross-sectional study among HIV-infected and uninfected Peruvian women aged ≥40. Dual X-ray absorptiometry was used to measure trunk and limb lean mass (LM) and fat mass (FM). Physical performance was assessed with the Short Physical Performance Battery (SPPB) and physical strength with a dynamometer. Sarcopenia was assessed based upon EWGSOP criteria. We used linear regression to model associations between BC, sarcopenia and physical scores.

**Results:** 104 HIV-infected and 212 uninfected women were enrolled with a mean age of 55.1±8.8 years. Among the HIV-infected group, years since diagnosis was 11.8±6 and all were on antiretroviral treatment. Mean SPPB score was 9.9 vs 10.8 (p<0.001) between both groups. Sarcopenia spectrum was found in 25.9% of HIV-infected and 23.1% of uninfected women. In multivariate regression analysis, trunk FM and older age were negatively correlated with physical performance among HIV-infected women. An increased percentage of severe sarcopenia was found among the HIV-infected, although our study was not specifically powered for this comparison.

**Conclusions:** HIV-infected women had significantly lower SPPB scores compared to women without HIV, and trunk FM and upper limb LM were independent predictors for the tests described. Prospective studies are needed in Latin America & the Caribbean to identify individuals at high risk for declines in physical function, and inform prevention guidelines.

## INTRODUCTION

With continued improvements in survival rates, management of chronic conditions and optimization of overall health has become a primary global health concern in the care of people living with HIV (PLWH)[1], particularly in lower-and-middle income countries where infrastructure for chronic disease management may be fragmented. Studies have shown that onset of aging-related comorbidities appears to occur in PLWH approximately a decade earlier compared with the general population[2]. The burden of aging in PLWH represents a relatively new and complex condition globally, that may lead to a decline in physical function and vulnerability to injury or developing other pathologies[3]. In Latin America & the Caribbean (LAC) few studies have focused on the impact of aging-related comorbidities among middle-aged and older PLWH. Therefore an important research agenda remains to characterize the epidemiology and outcomes of these co-morbidities taking into account sociocultural and health systems related factors unique to this region [4].

The musculoskeletal (MSK) system is significantly affected by HIV and its treatment. Compared with the general population, PLWH present with accelerated bone mineral density loss, increased incidence of low bone mass and heterogeneous disorders of body composition, often associated with metabolic consequences[5,6]. Alterations in body composition, such as sarcopenia and fat redistribution, can reflect important changes in bone and MSK health[7]. Consequently, sarcopenia has been described as an age-related decline in muscle mass that impairs physical function, with well-established associations with morbidity and mortality[8].

As PLWH age with these comorbidities, the heterogeneity of older individuals with HIV highlights the relevance of identifying those who are at greatest risk of premature aging[9]. The Short Physical Performance Battery (SPPB) is an objective physical functioning assessment that measures physical performance in older populations and has been extensively used in research settings to identify patients at risk of adverse events[10,11]. Physical function measures have multiple applications among aging adults including identification of those at increased likelihood of poor outcomes, which may allow clinicians to identity high-risk older individuals and prioritize them for preventive interventions[12].

Although more than half of PLWH worldwide currently are women, there is limited data about the specific characteristics of women aging with HIV[13]. Furthermore, no studies have focused on the relationship between body composition changes, sarcopenia and physical function among women with HIV in LAC. We aimed to explore the scope of this issue among Peruvian women aging with HIV using Dual X-ray absorptiometry (DXA)-based body composition measures and the well-established SPPB.

## METHODS

### Study design and participants

We conducted a hospital-based cross-sectional study. Between October 2019 and March 2020, participants were invited to participate at a large HIV-clinic from a public hospital in Lima, Peru. We enrolled HIV-positive women aged ≥40 years who presented for routine HIV-care visits during the study period, and concurrently recruited a control group of HIV-uninfected women aged ≥40 years. We used two strategies to recruit women in the control group, including inviting community-dwelling women from surrounding neighborhoods and as well as advertising the study among hospital staff (administrators, nurses and technicians). Exclusion criteria included pregnancy (assessed with urine pregnancy test at recruitment), previous diagnosis of or treatment for osteoporosis, and insufficient literacy to complete the questionnaire. Written informed consent was obtained in accordance with the procedures approved by the ethics committees from Yale School of Medicine, Universidad Peruana Cayetano Heredia and Hospital Nacional Arzobispo Loayza.

### Data collection and measures

Participants completed a self-administered questionnaire including socio-demographic and clinical data. Questionnaires were piloted before enrollment among 10 patients and 10 clinic staff to assure ease of use and clarity. HIV-related characteristics such as CD4^+^ T-cell count, viral load and antiretroviral therapy (ART) were obtained from medical records. HIV viral load suppression was defined as <50 copies/ml. Anthropometric measures were obtained using the same standardized stadiometer and weight scale for all participants. Waist and hip circumferences were measured to the closest 0.1 cm using a non-elastic tape measure.

### Body composition measurements

DXA was performed using a Hologic Discovery – WI 2009 series 84453 machine (Hologic Inc, Waltham, MA, USA). Scans of the whole body were performed using the automatic scan mode with participants wearing light clothing according to standard procedures. Body composition measures included trunk and limb lean mass (LM), fat mass (FM) and bone mineral content (BMC), all measured in grams. The fat mass index (FMI), skeletal muscle mass index (SMI) and appendicular skeletal muscle mass index (ASMI) were calculated as total body FM, total body LM and limb LM divided by the square of height (kg/m^2^), respectively[14]. Daily calibration and long-term DXA stability monitoring were performed according to standard protocols using manufacturer phantoms.

### Tests of physical function and strength

The SPPB was selected as an evidence-based, quick-to-perform physical function assessment. This tool has been used for more than 20 years to assess performance and functional status in aging populations[11]. The SPPB includes timed measures of standing balance in 3 positions (side-by-side, semi-tandem, and tandem), 4-meter gait speed, and time required to stand up and sit down 5 times. The test was performed according to the National Institute on Aging protocol[15]. Each measure was designated a score from 0 to 4, with 0 indicating inability to complete the test. Composite measures yielded a total score from 0 to 12, where a score ≤8 was used to indicate disability/low physical function[12,16].

Physical strength was assessed with hand grip strength using a JAMAR hydraulic hand dynamometer (Patterson Medical, IL, USA), which is the most widely used dynamometer in the research setting, and has been validated in several different patient populations and countries for measuring grip strength[17,18]. Force was measured in kilograms according to standardized procedures. Participants were asked to sit with their shoulder adducted and neutrally rotated, elbow flexed at 90° and with the forearm in a neutral position maximally squeeze the handle of the dynamometer with their dominant hand for three seconds. We set the dynamometer on the second handle position and performed the test three successive times, the average of the three trials was recorded.

### Sarcopenia Spectrum

Sarcopenia was defined and staged according to the European Sarcopenia Working Group (EWGSOP) guidelines[19], which recommends the presence of low muscle mass (ASMI≤5.67 kg/m^2^) and low muscle function (strength [grip strength≤20kg] or performance [SPPB≤8]) for diagnosis of this syndrome[20–22]. The “presarcopenia” stage was defined as low muscle mass without impact on muscle strength or performance. The “sarcopenia” stage was defined as low muscle mass, plus low muscle strength or physical performance and “severe sarcopenia” was defined when all three criteria of the definition were met.

### Statistical analysis

We described the sample characteristics and outcomes using standard frequency analysis, means, standard deviations, and proportions for all variables, as appropriate. Differences between HIV-infected and uninfected women were examined using independent t-test, x^2^, and Fisher exact test, as appropriate.

Unadjusted and adjusted linear regression models were adopted separately for the HIV-infected ad uninfected women to further analyze associations of independent variables (sociodemographic, clinical characteristics and body composition measurements) and physical function (SPPB and grip strength). Two multivariable models were constructed for the independent variables and SPPB, and grip strength separately. We first assessed for possible multicollinearity of the independent variables. Variables with a high correlation coefficient (r>0.40) were deemed collinear and excluded for the models. We fit the multivariate models using backward regression[23], starting with all the variables that showed a hypothesized relationship (p<0.10) in the bivariate model. We then removed non-significant (p>0.05) variables one at a time beginning with the least significant (largest p-value), in order to achieve the most parsimonious model.

Association between the sarcopenia spectrum (presarcopenia, sarcopenia and severe sarcopenia) and both sociodemographic and clinical related factors were explored with logistic regression analysis, applying the same steps mentioned above. Results were reported as unadjusted and adjusted standardized beta coefficients or odds ratio (OR) with 95% confidence intervals, as appropriate. All statistical analyses were performed using STATA version 16 (StataCorp, College Station, Texas, USA).

## RESULTS

### Sociodemographic and clinical characteristics

A total of 316 women were recruited in this study, including 104 HIV-infected and 212 uninfected women. The mean age of uninfected women was slightly older than women with HIV (56.4±8.8 vs 52.4±8.2 years, p<001). Women with HIV had a lower mean body mass index (BMI) (24.4 kg/m^2^ vs 27.6, p=0.05) compared to uninfected women, and a lower percentage of them had high school education or above (65.4% vs 85%, p<0.001) compared to the control group. Postmenopausal women represented 81% of the entire sample (Table 1).

**Table 1.** Sociodemographic and Clinical Characteristics by HIV status

|  | HIV-infected<br>N=104 | Uninfected<br>N=212 |
| --- | --- | --- |
| **Sociodemographic characteristics** |  |  |
| Age, mean (SD) | **52.38 (8.15)** | **56.37 (8.8)¥** |
| Marital status, n(%) |  |  |
| Single/divorced/separated/widowed, n(%) | **74 (71.15)** | **95 (44.81)¥** |
| Married/cohabitant, n(%) | 30 (28.85) | 117 (55.19) |
| Ethnicity |  |  |
| Caucasian, n(%) | 3 (2.88) | 24 (11.32) |
| Mestizo, n(%) | **97 (93.27)** | **185 (87.26)\*** |
| Black, n(%) | 4 (3.85) | 3 (1.42) |
| Education level |  |  |
| High school education or above, n(%) | **68 (65.38)** | **181 (85.38)¥** |
| **Clinical characteristics** |  |  |
| Body mass index (kg/m<sup>2</sup>), mean (SD) | **26.44 (5.1)** | **27.63 (4.12)\*** |
| Smoking ever, n (%) | 28 (26.92) | 39 (18.4) |
| Current alcohol use, n(%) | 25 (24.04) | 60 (28.3) |
| Current use of bone nutritional supplements, n(%) | 5 (4.81) | 11 (5.19) |
| Rheumatoid arthritis diagnosis, n(%) | - | 6 (2.83) |
| Postmenopausal status, n(%) | 80 (76.92) | 177 (83.49) |
| **HIV-related clinical data** |  |  |
| Years since HIV diagnosis mean (SD) | 11.81 (6.16) | - |
| Nadir CD4+ cell count mean (SD) | 264.32 (174.34) | - |
| Nadir HIV viral load mean (SD) | 128200.2 (275567.1) | - |
| Current CD4+ cell count mean (SD) | 593.23 (297.5) | - |
| Current HIV viral load mean (SD) | 30887.17 (222988.9) | - |
| AIDS diagnosis n(%) | 8 (7.69) | - |
| Duration ART exposure (years) mean (SD) | 9.86 (5.27) | - |
| PI exposure n(%) | 32 (30.77) | - |
| NRTI exposure n(%) | 103 (99.04) | - |
| **Body composition measurements** |  |  |
| Upper limb |  |  |
| FM (kg), mean (SD) | **3.34 (1.76)** | **3.79 (1.62)\*** |
| LM (kg), mean (SD) | 3.5 (0.78) | 3.53 (0.7) |
| BMC (kg), mean (SD) | **0.21 (0.04)** | **0.22 (0.04)\*** |
| Lower limb |  |  |
| FM (kg), mean (SD) | **6.53 (2.43)** | **8.14 (2.17)¥** |
| LM (kg), mean (SD) | 11.47 (2.18) | 11.47 (1.72) |
| BMC (kg), mean (SD) | **0.56 (0.12)** | **0.59 (0.11)\*** |
| Trunk FM (kg), mean (SD) | **11.54 (3.6)** | **12.71 (3.22)‡** |
| Total body |  |  |
| FM (kg), mean (SD) | **22.66 (7.24)** | **25.86 (6.51)¥** |
| LM (kg), mean (SD) | 37.33 (6.33) | 37.65 (4.88) |
| BMC (kg), mean (SD) | 1.74 (0.32) | 1.79 (0.28) |
| ASMI | 6.31 (1.04) | 6.3 (0.88) |
| FMI | **9.59 (3.02)** | **10.88 (2.67)¥** |
| SMI | 15.76 (2.42) | 15.84 (1.89) |

SD, standard deviation; FM, fat mass; LM, lean mass; BMC, bone mineral content; ASMI, appendicular skeletal muscle index; FMI, fat mass index; SMI, skeletal mass index; PI, protein inhibitors; NRTI, nucleoside reverse transcriptase inhibitors; CD4+ cell count = cell/mm<sup>3</sup>; HIV viral load = copies (ml); \*p ≤ 0.05; ‡p ≤ 0.01; ¥p ≤ 0.001

Among women with HIV, the mean time since HIV diagnosis was 11.8±6 years and all women were currently receiving ART (mean duration of treatment=9.9±5.3 years). The current mean CD4^+^ T-cell count was 593.2±297.5 cells/mm^3^ and the mean CD4^+^ T-cell count nadir was 264.3±174.3 cells/mm^3^. 78.8% had an undetectable viral load.

### Body composition, physical function and strength, and sarcopenia

In terms of body composition measures, women with HIV had decreased total FM (p<0.001) compared to uninfected women. Significant differences were found between FMI (p<0.001), upper limb FM (p=0.02), lower limb FM (p<0.001) and trunk FM (p=0.004) between both groups. LM was found to be lower in the upper limb and total body of HIV-infected women compared to the control group, but these comparisons were not statistically different.

The overall SPPB score was slightly lower in HIV-infected women compared to the control group (9.9 vs 10.8, p<0.001). A notable difference was observed in the proportion of patients with SPPB score ≤8, but this did not reach statistical significance (16.4% vs 4.7%, p=0.099) (Table 2). Small but statistically significant differences were found in SPPB sub-scores between the two groups, including the balance test scores (3.7 vs 3.9, p<0.001), gait speed test scores (3.6 vs 3,7, p=0.01) and chair stand test scores (2.8 vs 3.1, p<0.001). Average repeated grip strength was similar in the two groups (19.9±5.9 kg vs 19.8±5.4 kg).

**Table 2:**
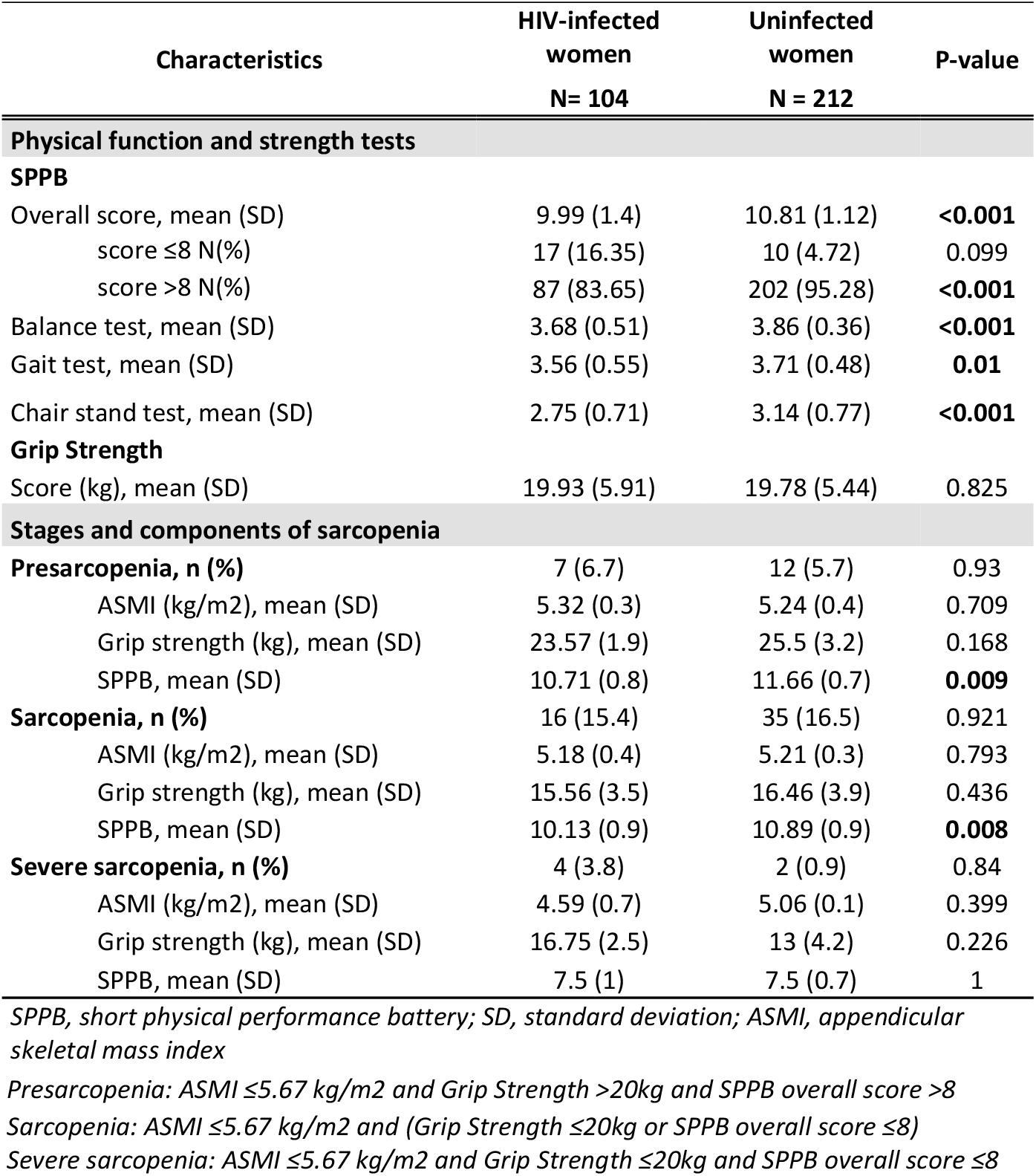
Physical function and strength test and Sarcopenia stages according to HIV status

In terms of the sarcopenia spectrum, 25.9% of HIV-infected and 23.1% uninfected women met criteria for either presarcopenia, sarcopenia or severe sarcopenia, but this difference did not reach statistical significance. Sarcopenia stage was the most common overall (15.4% in HIV-infected vs 16.5% uninfected women), and severe sarcopenia was higher in the HIV-infected subset (3.8% vs 0.9%), but was not statistically different. Moreover, although ASMI was higher among uninfected women compared with HIV-infected women for both sarcopenia and severe sarcopenia stages, the differences were not statically significant. **(Table 2)**

### Associations between clinical characteristics, body composition, and physical function assessments

In terms of physical function, the multivariable analysis among uninfected women showed that higher SPPB score was independently associated with current alcohol use (p=0.01) and higher lower limb BMC (p=0.03), and lower SPPB score was significantly associated with older age (p<0.001) and higher BMI (p<0.001) (Table 3). However, in the multivariable model for HIV-infected women, only trunk FM (p=0.038) and older age (p=0.002) were negatively correlated with SPPB score. (Table 4)

**Table 3.** Unadjusted and Adjusted linear regression for SPPB and Grip Strength, Uninfected Women (N=212)

| Independent variables | SPPB |  |  |  | Grip strength test |  |  |  |
| --- | --- | --- | --- | --- | --- | --- | --- | --- |
|  | Unadjusted |  | Adjusted |  | Unadjusted |  | Adjusted |  |
|  | B | 95% C.I. | B | 95% C.I. | B | 95% C.I. | B | 95% C.I. |
| **Body composition measurements** |  |  |  |  |  |  |  |  |
| Trunk FM | **-0.064** | **-0.117 to -0.012\*** | - | - | 0.147 | -0.082 to 0.375 | - | - |
| Trunk BMC | **1.892** | **0.001 to 3.784\*** | - | - | **15.208** | **7.281 to 23.135¥** | - | - |
| Trunk LM | 0.002 | -0.064 to 0.067 | - | - | **0.386** | **0.110 to 0.662‡** | - | - |
| Trunk %fat | **-0.056** | **-0.093 to -0.019‡** | - | - | -0.046 | -0.208 to 0.116 | - | - |
| Lower limb FM | -0.056 | -0.135 to 0.023 | - | - | 0.274 | -0.065 to 0.612 | - | - |
| Lower limb BMC | **1.998** | **0.427 to 3.569\*** | **1.551** | **0.165 to 2.938\*** | **15.495** | **8.985 to 22.004¥** | - | - |
| Lower limb LM | 0.024 | -0.076 to 0.124 | - | - | **0.818** | **0.403 to 1.234¥** | - | - |
| Lower limb %fat | -0.023 | -0.056 to 0.010 | - | - | -0.041 | -0.183 to 0.100 | - | - |
| Upper limb FM | **-0.151** | **-0.255 to -0.046‡** | - | - | 0.026 | -0.431 to 0.482 | - | - |
| Upper limb BMC | 3.730 | -0.188 to 7.648 | - | - | **41.255** | **25.254 to 57.256¥** | - | - |
| Upper limb LM | 0.024 | -0.076 to 0.124 | - | - | **1.746** | **0.713 to 2.779¥** | **1.804** | **0.997 to 2.611¥** |
| Upper limb %fat | **-0.029** | **-0.052 to -0.005\*** | - | - | -0.097 | -0.197 to 0.003 | - | - |
| Total body FM | **-0.032** | **-0.058 to -0.006\*** | - | - | 0.072 | -0.041 to 0.186 | - | - |
| Total body BMC | **0.831** | **0.234 to 1.427‡** | - | - | **5.776** | **3.290 to 8.263¥** | - | - |
| Total body LM | 0.002 | -0.033 to 0.038 | - | - | **0.272** | **0.125 to 0.419¥** | - | - |
| Total body %fat | **-0.055** | **-0.094 to -0.017‡** | - | - | -0.072 | -0.242 to 0.097 | - | - |
| **Sociodemographic and clinical characteristics** |  |  |  |  |  |  |  |  |
| Age | **-0.067** | **-0.084 to -0.050¥** | **-0.043** | **-0.060 to -0.026¥** | **-0.194** | **-0.273 to -0.114¥** | **-0.161** | **-0.227 to -0.094¥** |
| BMI | **-0.061** | **-0.102 to -0.020‡** | **-0.059** | **-0.092 to -0.026¥** | 0.105 | -0.074 to 0.284 | - | - |
| Smoking ever | 0.378 | -0.063 to 0.819 | - | - | 0.578 | -1.326 to 2.482 | - | - |
| Current alcohol use | **0.488** | **0.112 to 0.864\*** | **0.406** | **0.090 to 0.723\*** | -0.064 | -1.703 to 1.575 | - | - |
| Rheumatoid arthritis diagnosis | **-1.767** | **-2.776 to -0.758¥** | - | - | -3.895 | -8.316 to 0.526 | - | - |
| Menopause status | **-0.750** | **-1.202 to -0.298¥** | - | - | **-3.338** | **-5.274 to -1.402¥** | - | - |
| Bone nutritional supplements, ever | -0.085 | -0.861 to 0.690 | - | - | -0.827 | -4.154 to 2.500 | - | - |

SPPB, short physical performance battery; FM, fat mass; LM, lean mass; BMC, bone mineral content; BMI, body mass index; %fat, percentage of FM; \* $p \leq 0.05$ ; ‡ $p \leq 0.01$ ; ¥ $p \leq 0.001$

**Table 4.**
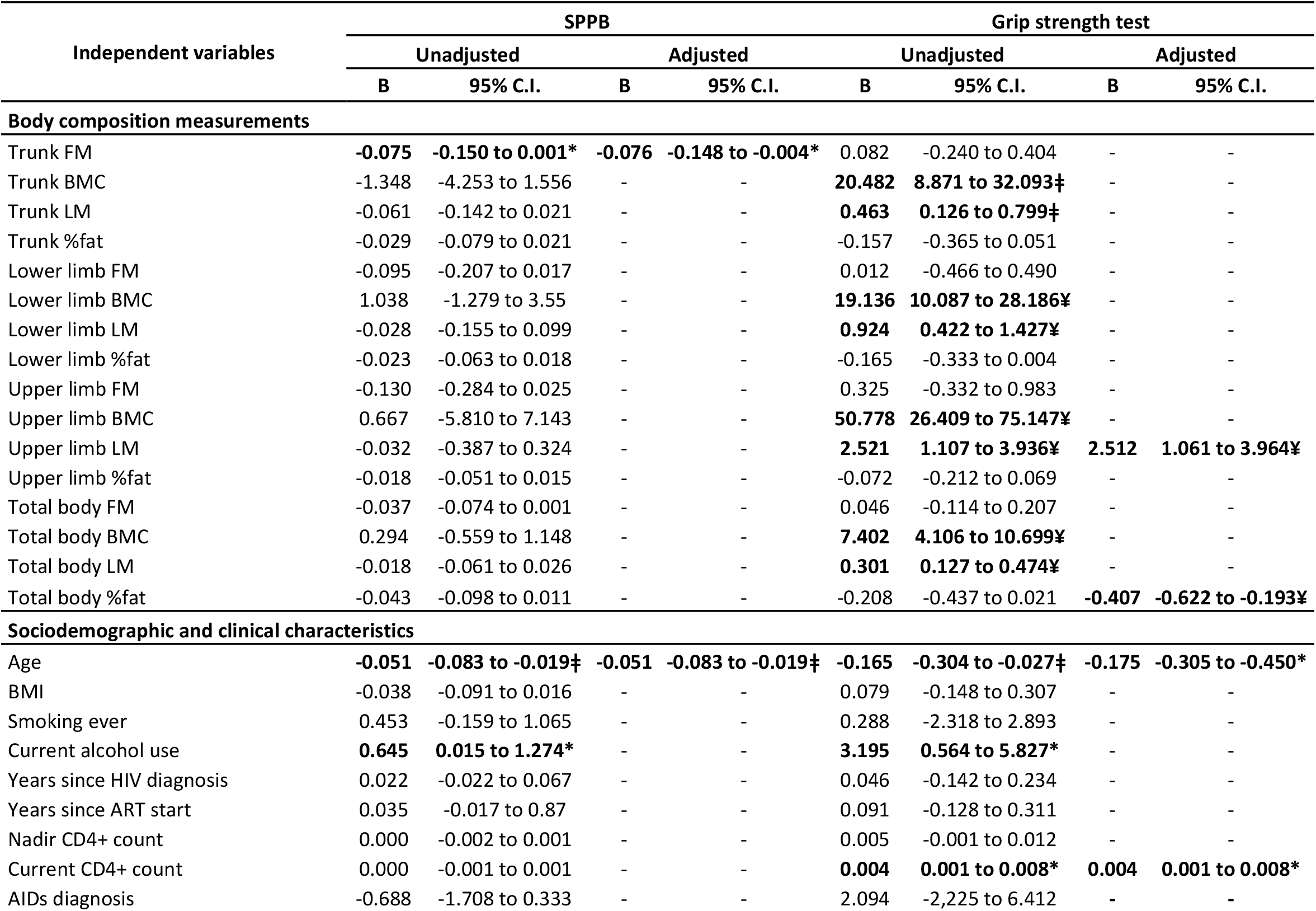
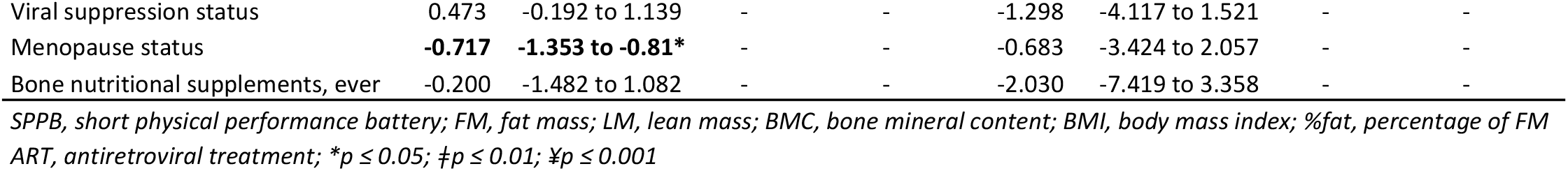
Unadjusted and Adjusted linear regression for SPPB and Grip Strength, HIV-infected women (N=104)

Regarding physical strength, in the uninfected women multivariable model, greater grip strength was positively correlated with upper limb LM (p<0.001), and negatively correlated with older age (p<0.001) (Table 3). Similarly, among HIV-infected women, the multivariable model showed upper limb LM (p<0.001) and higher CD4^+^ T-cell count (p=0.012) were positively correlated with grip strength, and older age (p=0.13) and total body fat percentage (p<0.001) were negatively correlated with this outcome. (Table 4)

### Association between clinical characteristics and sarcopenia

Significant associations were found among the HIV-infected univariate model in terms of marital status, where sarcopenia risk reduced with being married/cohabitant (OR 0.14, p=0.01). Higher BMI (OR 0.57, p<0.001) and higher current CD4^+^ T-cell count (OR 0.99, p<0.5) were also significantly associated with reduced sarcopenia risk, and history of AIDS (OR 10.71, p<0.01) and protease inhibitor (PI) exposure (OR 3.53, p<0.01) were associated with increased sarcopenia risk. Likewise, in the multivariable model, higher BMI (OR 0.56, p<0.001) and being married/cohabitant (OR 0.10, p<0.5) were associated with reduced sarcopenia risk, along with PI exposure (OR 0.19, p<0.01) (Table 5).

**Table 5.**
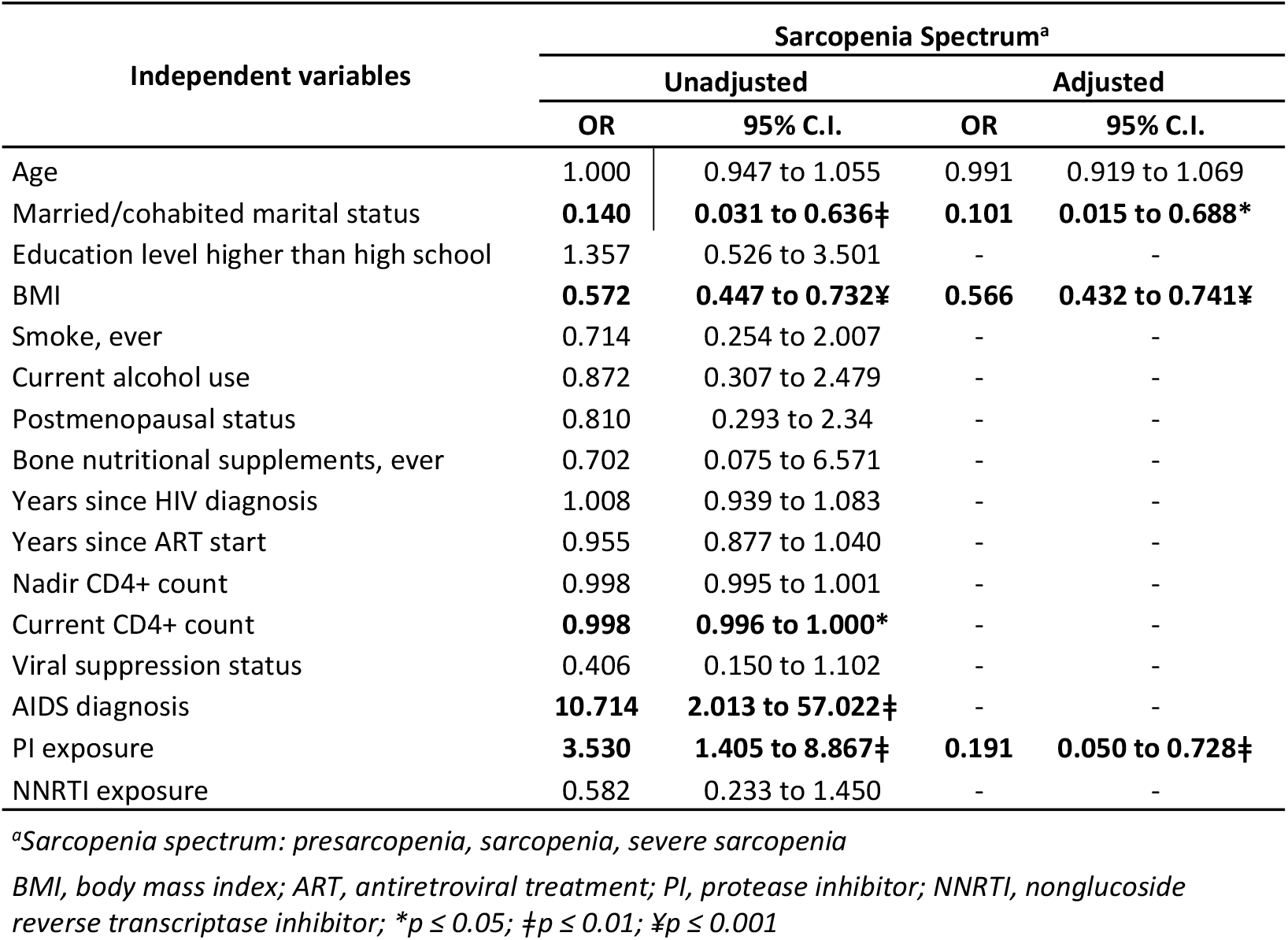
Unadjusted and Adjusted logistic regression for Sarcopenia Spectrum, HIV-infected women (N=104)

| Independent variables | Sarcopenia Spectrum<sup>a</sup> |  |  |  |
| --- | --- | --- | --- | --- |
|  | Unadjusted |  | Adjusted |  |
|  | OR | 95% C.I. | OR | 95% C.I. |
| Age | 1.000 | 0.947 to 1.055 | 0.991 | 0.919 to 1.069 |
| Married/cohabited marital status | **0.140** | **0.031 to 0.636‡** | **0.101** | **0.015 to 0.688\*** |
| Education level higher than high school | 1.357 | 0.526 to 3.501 | - | - |
| BMI | **0.572** | **0.447 to 0.732¥** | **0.566** | **0.432 to 0.741¥** |
| Smoke, ever | 0.714 | 0.254 to 2.007 | - | - |
| Current alcohol use | 0.872 | 0.307 to 2.479 | - | - |
| Postmenopausal status | 0.810 | 0.293 to 2.34 | - | - |
| Bone nutritional supplements, ever | 0.702 | 0.075 to 6.571 | - | - |
| Years since HIV diagnosis | 1.008 | 0.939 to 1.083 | - | - |
| Years since ART start | 0.955 | 0.877 to 1.040 | - | - |
| Nadir CD4+ count | 0.998 | 0.995 to 1.001 | - | - |
| Current CD4+ count | **0.998** | **0.996 to 1.000\*** | - | - |
| Viral suppression status | 0.406 | 0.150 to 1.102 | - | - |
| AIDS diagnosis | **10.714** | **2.013 to 57.022‡** | - | - |
| PI exposure | **3.530** | **1.405 to 8.867‡** | **0.191** | **0.050 to 0.728‡** |
| NNRTI exposure | 0.582 | 0.233 to 1.450 | - | - |

<sup>a</sup>Sarcopenia spectrum: presarcopenia, sarcopenia, severe sarcopenia
BMI, body mass index; ART, antiretroviral treatment; PI, protease inhibitor; NNRTI, nonglucoside reverse transcriptase inhibitor; \* $p \leq 0.05$ ; ‡ $p \leq 0.01$ ; ¥ $p \leq 0.001$

## DISCUSSION

Recognition of aging-related concerns among PLWH, including physical function and sarcopenia, is key to distinguishing individuals at greatest risk for downstream musculoskeletal complications[10]. Our study is the first to explore the relationship between body composition alterations, sarcopenia and physical function among women aging with and without HIV in LAC using well-established assessment tools. HIV-infected women had significantly lower SPPB scores compared to uninfected women and an increased percentage of severe sarcopenia was found in the HIV-infected group.

HIV epidemiology in Peru has changed significantly over the past decade. From 2014 to 2018, the rate of viral suppression among PLWH receiving ART rose from 36% to 65%, due to widespread free antiretroviral access through health clinics around Peru[24],[25]. Consistent with this trend, 78.8% of our HIV-positive sample had an undetectable viral load. While improved viral control leads to improved survival among PLWH, data regarding this epidemiological transition among women or men aging with HIV in Peru is completely lacking, and is quite limited from other countries in the LAC region [26],[27].

Changes in body composition, such as fat redistribution, have been shown to impact physical function and risk for mortality[1]. Since 2013, the International Society for Clinical Densitometry has included ART use in PLWH as one of the indications for using DXA-based body measurements of body composition, in particular for patients receiving agents associated with risk of lipoatrophy [28,29]. Among these changes, peripheral (face and limbs) fat atrophy is commonly reported[30,31]. In our study, women with HIV had lower FM in the limbs and trunk compared to uninfected women, a finding that was statistically significant. Prevalence of fat redistribution varies widely across reports (7-65%) in low-and middle-income countries (LMICs)[32], and in LAC, only one study from Brazil has assessed body composition changes in PLWH with objective measurements such as DXA scan. Among 262 PLWH (43% women) the authors found a 40.8% overall prevalence of lipodystrophy based upon DXA-derived fat mass ratio, underscoring the importance of early screening and intervention in this population to prevent complications[33]. Interestingly, in the Brazilian sample, prevalence of lipodystrophy measured by DXA among women was only 15.8%, compared to 48.8% among men (p<0.05), however the authors attributed this finding to their use of a cutoff for lipodystrophy that was previously standardized in men, but not women, as the prevalence of lipodystrophy among women measured by clinical exam was 56.5% in women, versus 47.7% in men.

In terms of impact of these anthropometric changes on physical performance of PLWH, studies conducted in upper-income countries have reported that HIV-infected individuals appear to have worse physical function compared to those without HIV[34–36]. Among participants in our study, women with HIV had a modestly but significantly lower SPPB score compared to the control group. Compared to another study of 176 PLWH (19% women) in the United States (US) with mean age of 54.6 years, both HIV-infected and infected women of our sample had lower SPPB scores[12]. Additionally, 16.4% of HIV-infected and 4% of uninfected women in our study had a SPPB score ≤8 points, which has been shown to be predictive for subsequent disability and mortality[16,37]. A similar study among 65 middle-aged and older (45-55 years) women with HIV in the US with well-controlled disease (mean current CD4^+^ T-cell 675 cells/mm3, 89% viral load <75 copies/mL) found that 20% of women met criteria for low physical function based upon the SPPB[38]. Even though in our study population, HIV-related biomarkers were relatively stable (mean CD4^+^ 593.3 cell/mm^3^; 78.4% viral suppression), a considerable difference was detected in terms of proportion of women with SPPB score ≤8 in the HIV-infected versus uninfected groups. Accordingly, one of the largest studies to apply the SPPB in PLWH found that having well-controlled CD4^+^ T-cell counts and viral load moderated, but did not eliminate, the odds of reduced physical performance compared the general population[10].

In our study, older age was negatively associated with SPPB score in both HIV-infected and uninfected women. These findings are consistent with the scarce literature conducted mostly in upper-income countries, and to a lesser extent, Africa[12,38,39].Furthermore, trunk FM was negatively associated with SPPB in the multivariable analyses for both HIV-infected and uninfected women. These results are similar to those reported in previous studies in different aging populations[40–43], however, none of them included HIV-infected individuals.

In terms of physical strength, upper limb LM was positively associated with grip strength in regression models for both HIV-infected and uninfected individuals, consistent with previous evidence describing muscle mass as an important predictor of physical function[43]. In our study, although total body LM was slightly higher in uninfected women, differences were not significant. Moreover, among the HIV-infected multivariate model, total body fat percentage was negatively correlated with grip strength, which is also in line with the negative relationship observed in our study between trunk FM and SPPB scores. The mechanisms and natural history of altered lean mass among aging PLWH receiving ART remain poorly understood. Grant et al. described a “return to health effect” where PLWH gained more LM during the first 96 weeks of ART than HIV-uninfected individuals as their immune function and overall health status improves with treatment. However, after that period, patients with HIV had a decrease in LM despite virologic suppression[1]. Other studies have demonstrated a correlation between elevated proinflammatory cytokine levels (e.g. interleukin-6 and C-reactive protein) with low lean mass[44].

Lean or muscle mass is another body compartment frequently affected by HIV[1]. To date, studies on sarcopenia among PLWH are limited and have applied different operational definitions[8]. To our knowledge, ours is the first study that reports sarcopenia among Peruvian women with HIV using the EWGSOP recommendations. The rationale behind this multicomponent definition, which takes into account muscle mass, function and strength, is that characterizing sarcopenia simply in terms of muscle mass significantly limits the clinical utility of the definition[45]. In our study, the majority of women that presented with this syndrome were classified as “sarcopenia” stage. Likewise, a recent metanalysis across 13 studies of PLWH (range: 35-60 years of age) from different geographic regions, including two studies from Brazil, found a pooled sarcopenia prevalence of 13.2% (CI=5.2-22.9%) using two operational definitions. These included (1) the presence of low LM (four studies) measured by DXA or bioimpedance analysis, or (2) the EWGSOP definition (nine studies)[46]. Our multivariable regression model found that BMI, protease inhibitor (PI) exposure and being married/cohabitant were negatively associated with a sarcopenia spectrum diagnosis. Some of these associations are consistent with previous reports, where higher BMI was associated with a lower odds of sarcopenia[46]. However, although PI exposure was positively associated with sarcopenia in the unadjusted analysis, in the multivariable model it was a protective factor, which contradicts previous data demonstrating increased risk of body composition and metabolic alterations[47,48]. Moreover, in our sample of patients, the proportion of “severe sarcopenia” was higher in the HIV-infected women compared to uninfected group (3.8% vs 0.9%), however this difference did not achieve statistical significance perhaps due to the limited sample size. This finding may suggest worse prognosis for HIV-positive individuals and warrants further evaluation in larger-scale, longitudinal studies.

This study has some important limitations. First, as this was a single-center study our findings might not be generalizable to all women with HIV in Peru, nor to those outside Peru due to differences in cultural and environmental factors. However, this study was conducted in one of the largest public HIV clinics in Peru’s capital city. Second, the cross-sectional nature of our study means that associations between physical strength, performance measures, and sarcopenia spectrum outcomes cannot be inferred to suggest causality, furthermore, we didn’t carry out a power calculation, limiting the statistical power of this study. However, our goal was to explore possible relationships between body composition and physical function to further promote future studies in this topic among Latin American populations. Finally, the EWGSOP published updated guidelines in 2019[49] that propose a new algorithm for screening and diagnosing sarcopenia, which incorporates the SARC-F screening tool and a slightly modified sequence of the sarcopenia components used in the original guidelines (LM, muscle strength and physical performance). As this new algorithm utilizes European region-specific cutoff thresholds which have not yet been adapted to Latin American populations (whereas in certain regions such as Asia, corresponding guidelines have already been published with thresholds specifically to Asian populations[50]), we opted to apply the well-established 2010 EWGSOP guidelines which have been utilized previously in LAC populations[51]. Future studies are needed to test and develop appropriate threshold for the new EWGSOP algorithm among individuals from LAC.

In summary, as HIV increasingly shifts to a chronic condition, and the proportion of patients aging with HIV globally continues to grow, research characterizing the effect of aging-related comorbidities among this population using well-validated tools is critically needed[52]. Much remains unknown regarding the extent to which alterations in body composition impact physical function and sarcopenia among women with HIV. We found that HIV-infected women in our study had significantly lower SPPB scores compared to uninfected women, and that trunk FM and upper limb LM were independent predictors for physical function. Additionally, a higher prevalence of severe sarcopenia was observed among HIV-infected women in our study. These findings underscore the necessity for larger prospective studies among PLWH, particularly in LAC, to help identify individuals at greatest risk for declines in physical function and sarcopenia. Our data suggest that targeting lower trunk/abdominal fat mass and increasing limb lean mass could potentially have a positive impact on physical function outcomes. However, intervention studies are needed to inform clinical practice guidelines with regards to optimizing musculoskeletal health among women with HIV.

## Data Availability

All dataset from this project is available for data transparency purposes on https://osf.io/qtyv9/?view_only=b2a8a2cf2e73488ba7b6fde07f30e103

## Acknowledgments

We thank Debbie Miyasato, Eduardo Matos, Ana Maria Guerrero, Maricruz Huaman, Karim Sanchez, Rocio Galvez and Miriam Santos from the HIV clinic and Radiology Department at Hospital Nacional Arzobispo Loayza for their invaluable help during data collection. Additionally, we greatly thank Cesar Cárcamo for his important input regarding data analysis.

## DECLARATIONS

### Ethics approval

This study was reviewed and approved by the institutional review boards of Yale School of Medicine, Universidad Peruana Cayetano Heredia and Hospital Nacional Arzobispo Loayza. The procedures used in this study adhere to the tenets of the Declaration of Helsinki.

### Consent to participate

All willing participants provided written informed consent after a comprehensive explanation of the study procedures

### Consent for publication

All participants enrolled provided consent for publication of de-identified data in journal article

### Availability of data and material

All data and material are available for data transparency purposes and are available from corresponding author on reasonable request

### Code availability

All statistical code is available for data transparency purposes

## Authors contributions

Diego M. Cabrera, Evelyn Hsieh and Patricia J. Garcia contribute to study conception, design, material preparation, data collection and analysis, manuscript preparation, review and editing. Mijahil P. Cornejo and Yvett Pinedo were involved in implementation, data collection and manuscript review. All authors read and approved the final manuscript.

## REFERENCES

1. Grant PM, Kitch D, McComsey GA, Collier AC, Bartali B, Koletar SL, et al. Long-term body composition changes in antiretroviral-treated HIV-infected individuals. AIDS. 2016;30: 2805–2813. doi:10.1097/QAD.0000000000001248

2. Guaraldi G, Orlando G, Zona S, Menozzi M, Carli F, Garlassi E, et al. Premature age-related comorbidities among HIV-infected persons compared with the general population. Clin Infect Dis. 2011;53: 1120–1126. doi:10.1093/cid/cir627

3. Nasi M, De Biasi S, Gibellini L, Bianchini E, Pecorini S, Bacca V, et al. Ageing and inflammation in patients with HIV infection. Clin Exp Immunol. 2017;187: 44–52. doi:10.1111/cei.12814

4. Cabrera DM, Diaz MM, Grimshaw A, Salvatierra J, Garcia PJ, Hsieh E. Aging with HIV in Latin America and the Caribbean: a Systematic Review. Curr HIV/AIDS Rep. 2021;18: 1–47. doi:10.1007/s11904-020-00538-7

5. Tebas P, Powderly WG, Claxton S, Marin D, Tantisiriwat W, Teitelbaum SL, et al. Accelerated bone mineral loss in HIV-infected patients receiving potent antiretroviral therapy: AIDS. 2000;14: F63–F67. doi:10.1097/00002030-200003100-00005

6. Sacilotto LB, Pereira PCM, Manechini JPV, Papini SJ. Body Composition and Metabolic Syndrome Components on Lipodystrophy Different Subtypes Associated with HIV. J Nutr Metab. 2017;2017: 8260867. doi:10.1155/2017/8260867

7. Sharma A, Tian F, Yin MT, Keller MJ, Cohen M, Tien PC. Association of Regional Body Composition with Bone Mineral Density in HIV-infected and Uninfected Women: Women’s Interagency HIV Study. J Acquir Immune Defic Syndr. 2012;61: 469–476. doi:10.1097/QAI.0b013e31826cba6c

8. Abdul Aziz SA, Mcstea M, Ahmad Bashah NS, Chong ML, Ponnampalavanar S, Syed Omar SF, et al. Assessment of sarcopenia in virally suppressed HIV-infected Asians receiving treatment. AIDS. 2018;32: 1025–1034. doi:10.1097/QAD.0000000000001798

9. Brañas F, Jiménez Z, Sánchez-Conde M, Dronda F, López-Bernaldo De Quirós JC, Pérez-Elías MJ, et al. Frailty and physical function in older HIV-infected adults. Age and Ageing. 2017;46: 522–526. doi:10.1093/ageing/afx013

10. Greene M, Covinsky K, Astemborski J, Piggott DA, Brown T, Leng S, et al. The relationship of physical performance with HIV disease and mortality. AIDS. 2014;28: 2711–2719. doi:10.1097/QAD.0000000000000507

11. Guralnik JM, Simonsick EM, Ferrucci L, Glynn RJ, Berkman LF, Blazer DG, et al. A short physical performance battery assessing lower extremity function: association with self-reported disability and prediction of mortality and nursing home admission. J Gerontol. 1994;49: M85–94. doi:10.1093/geronj/49.2.m85

12. Crane HM, Miller ME, Pierce J, Willig AL, Case ML, Wilkin AM, et al. Physical Functioning Among Patients Aging With Human Immunodeficiency Virus (HIV) Versus HIV Uninfected: Feasibility of Using the Short Physical Performance Battery in Clinical Care of People Living With HIV Aged 50 or Older. Open Forum Infect Dis. 2019;6. doi:10.1093/ofid/ofz038

13. Brañas F, Sánchez-Conde M, Carli F, Menozzi M, Raimondi A, Milic J, et al. Sex Differences in People Aging With HIV: JAIDS Journal of Acquired Immune Deficiency Syndromes. 2020;83: 284–291. doi:10.1097/QAI.0000000000002259

14. Hou Y, Xie Z, Zhao X, Yuan Y, Dou P, Wang Z. Appendicular skeletal muscle mass: A more sensitive biomarker of disease severity than BMI in adults with mitochondrial diseases. PLoS One. 2019;14. doi:10.1371/journal.pone.0219628

15. Short Physical Performance Battery (SPPB). In: National Institute on Aging [Internet]. [cited 6 Jun 2020]. Available: https://www.nia.nih.gov/research/labs/leps/short-physical-performance-battery-sppb

16. Erlandson KM, Allshouse AA, Jankowski CM, Duong S, MaWhinney S, Kohrt WM, et al. A Comparison of Functional Status Instruments in HIV-Infected Adults on Effective Antiretroviral Therapy. HIV Clin Trials. 2012;13: 324–334. doi:10.1310/hct1306-324

17. Alahmari KA, Silvian SP, Reddy RS, Kakaraparthi VN, Ahmad I, Alam MM. Hand grip strength determination for healthy males in Saudi Arabia: A study of the relationship with age, body mass index, hand length and forearm circumference using a hand-held dynamometer. J Int Med Res. 2017;45: 540–548. doi:10.1177/0300060516688976

18. Leong DP, Teo KK, Rangarajan S, Kutty VR, Lanas F, Hui C, et al. Reference ranges of handgrip strength from 125,462 healthy adults in 21 countries: a prospective urban rural epidemiologic (PURE) study. J Cachexia Sarcopenia Muscle. 2016;7: 535–546. doi:10.1002/jcsm.12112

19. Cruz-Jentoft AJ, Baeyens JP, Bauer JM, Boirie Y, Cederholm T, Landi F, et al. Sarcopenia: European consensus on definition and diagnosis. Age Ageing. 2010;39: 412–423. doi:10.1093/ageing/afq034

20. Baumgartner RN, Koehler KM, Gallagher D, Romero L, Heymsfield SB, Ross RR, et al. Epidemiology of sarcopenia among the elderly in New Mexico. Am J Epidemiol. 1998;147: 755–763. doi:10.1093/oxfordjournals.aje.a009520

21. Lauretani F, Russo CR, Bandinelli S, Bartali B, Cavazzini C, Di Iorio A, et al. Age-associated changes in skeletal muscles and their effect on mobility: an operational diagnosis of sarcopenia. J Appl Physiol. 2003;95: 1851–1860. doi:10.1152/japplphysiol.00246.2003

22. Guralnik JM, Ferrucci L, Pieper CF, Leveille SG, Markides KS, Ostir GV, et al. Lower extremity function and subsequent disability: consistency across studies, predictive models, and value of gait speed alone compared with the short physical performance battery. J Gerontol A Biol Sci Med Sci. 2000;55: M221–231. doi:10.1093/gerona/55.4.m221

23. Sauerbrei W, Royston P, Binder H. Selection of important variables and determination of functional form for continuous predictors in multivariable model building. Stat Med. 2007;26: 5512–5528. doi:10.1002/sim.3148

24. Cáceres C. Estudio sobre el Continuo de Atención de las Personas con VIH. Lima, Perú: Centro de Investigación Interdisciplinaria en sexualidad, SIDA y Sociedad; 2019 p. 71. Report No.: 8.

25. Garcia-Fernandez L, Novoa R, Huaman B, Benites C. Continuo de la atención de personas que viven con VIH y brechas para el logro de las metas 90-90-90 en Perú. Revista Peruana de Medicina Experimental y Salud Publica. 2018;35: 491–496. doi:10.17843/rpmesp.2018.353.3853

26. Cardoso SW, Torres TS, Santini-Oliveira M, Marins LMS, Veloso VG, Grinsztejn B. Aging with HIV: a practical review. The Brazilian Journal of Infectious Diseases. 2013;17: 464–479. doi:10.1016/j.bjid.2012.11.007

27. Crabtree-Ramirez B, Del Rio C, Grinsztejn B, Sierra-Madero J. HIV and noncommunicable diseases (NCDs) in Latin America: A call for an integrated and comprehensive response. Journal of Acquired Immune Deficiency Syndromes. 2014;67: S96–S98. doi:10.1097/QAI.0000000000000261

28. Shepherd JA, Baim S, Bilezikian JP, Schousboe JT. Executive summary of the 2013 International Society for Clinical Densitometry Position Development Conference on Body Composition. J Clin Densitom. 2013;16: 489–495. doi:10.1016/j.jocd.2013.08.005

29. Shuhart CR, Yeap SS, Anderson PA, Jankowski LG, Lewiecki EM, Morse LR, et al. Executive Summary of the 2019 ISCD Position Development Conference on Monitoring Treatment, DXA Cross-calibration and Least Significant Change, Spinal Cord Injury, Peri-prosthetic and Orthopedic Bone Health, Transgender Medicine, and Pediatrics. J Clin Densitom. 2019;22: 453–471. doi:10.1016/j.jocd.2019.07.001

30. Chiţu-Tişu CE, Barbu EC, Lazăr M, Bojincă M, Tudor A-M, Hristea A, et al. Body composition in HIV-infected patients receiving highly active antiretroviral therapy. Acta Clinica Belgica. 2017;72: 55–62. doi:10.1080/17843286.2016.1240426

31. Nuvoli S, Caruana G, Babudieri S, Solinas P, Pellicanò G, Piras B, et al. Body fat changes in HIV patients on highly active antiretroviral therapy (HAART): a longitudinal DEXA study. European Review for Medical and Pharmacological Sciences. 2018;22: 1852–1859. doi:10.26355/eurrev_201803_14606

32. Ali MK, Magee MJ, Dave JA, Ofotokun I, Tungsiripat M, Jones TK, et al. HIV and metabolic, body, and bone disorders: what we know from low- and middle-income countries. J Acquir Immune Defic Syndr. 2014;67 Suppl 1: S27–39. doi:10.1097/QAI.0000000000000256

33. Beraldo RA, Santos AP dos, Guimarães MP, Vassimon HS, Paula FJA de, Machado DRL, et al. Redistribuição de gordura corporal e alterações no metabolismo de lipídeos e glicose em pessoas vivendo com HIV/AIDS. Revista Brasileira de Epidemiologia. 2017;20: 526–536. doi:10.1590/1980-5497201700030014

34. Terzian AS, Holman S, Nathwani N, Robison E, Weber K, Young M, et al. Factors associated with preclinical disability and frailty among HIV-infected and HIV-uninfected women in the era of cART. J Womens Health (Larchmt). 2009;18: 1965–1974. doi:10.1089/jwh.2008.1090

35. Desquilbet L, Jacobson LP, Fried LP, Phair JP, Jamieson BD, Holloway M, et al. HIV-1 Infection Is Associated With an Earlier Occurrence of a Phenotype Related to Frailty. J Gerontol A Biol Sci Med Sci. 2007;62: 1279–1286. doi:10.1093/gerona/62.11.1279

36. Piggott DA, Muzaale AD, Mehta SH, Brown TT, Patel KV, Leng SX, et al. Frailty, HIV infection, and mortality in an aging cohort of injection drug users. PLoS ONE. 2013;8: e54910. doi:10.1371/journal.pone.0054910

37. Pavasini R, Guralnik J, Brown JC, di Bari M, Cesari M, Landi F, et al. Short Physical Performance Battery and all-cause mortality: systematic review and meta-analysis. BMC Med. 2016;14. doi:10.1186/s12916-016-0763-7

38. Baranoski AS, Harris A, Michaels D, Miciek R, Storer T, Sebastiani P, et al. Relationship Between Poor Physical Function, Inflammatory Markers, and Comorbidities in HIV-Infected Women on Antiretroviral Therapy. J Womens Health (Larchmt). 2014;23: 69–76. doi:10.1089/jwh.2013.4367

39. Lwanga I, Nabaggala MS, Kiragga A, Calcagno A, Guaraldi G, Lamorde M, et al. Implementing routine physical function screening among elderly HIV-positive patients in Uganda. AIDS Care. 2019; 1–4. doi:10.1080/09540121.2019.1703888

40. Kim JH, Chon J, Soh Y, Han YR, Won CW, Lee SA. Trunk fat mass correlates with balance and physical performance in a community-dwelling elderly population: Results from the Korean Frailty and aging cohort study. Medicine. 2020;99: e19245. doi:10.1097/MD.0000000000019245

41. Hicks GE, Simonsick EM, Harris TB, Newman AB, Weiner DK, Nevitt MA, et al. Trunk muscle composition as a predictor of reduced functional capacity in the health, aging and body composition study: the moderating role of back pain. J Gerontol A Biol Sci Med Sci. 2005;60: 1420–1424. doi:10.1093/gerona/60.11.1420

42. Woo J, Leung J, Kwok T. BMI, body composition, and physical functioning in older adults. Obesity (Silver Spring). 2007;15: 1886–1894. doi:10.1038/oby.2007.223

43. Valentine RJ, Misic MM, Rosengren KS, Woods JA, Evans EM. Sex impacts the relation between body composition and physical function in older adults. Menopause. 2009;16: 518–523. doi:10.1097/gme.0b013e31818c931f

44. PrayGod G, Blevins M, Woodd S, Rehman AM, Jeremiah K, Friis H, et al. A longitudinal study of systemic inflammation and recovery of lean body mass among malnourished HIV-infected adults starting antiretroviral therapy in Tanzania and Zambia. Eur J Clin Nutr. 2016;70: 499–504. doi:10.1038/ejcn.2015.221

45. Goodpaster BH, Park SW, Harris TB, Kritchevsky SB, Nevitt M, Schwartz AV, et al. The loss of skeletal muscle strength, mass, and quality in older adults: the health, aging and body composition study. J Gerontol A Biol Sci Med Sci. 2006;61: 1059–1064. doi:10.1093/gerona/61.10.1059

46. Oliveira VHF, Borsari AL, Webel AR, Erlandson KM, Deminice R. Sarcopenia in people living with the Human Immunodeficiency Virus: a systematic review and meta-analysis. Eur J Clin Nutr. 2020;74: 1009–1021. doi:10.1038/s41430-020-0637-0

47. van der Valk M, Gisolf EH, Reiss P, Wit FW, Japour A, Weverling GJ, et al. Increased risk of lipodystrophy when nucleoside analogue reverse transcriptase inhibitors are included with protease inhibitors in the treatment of HIV-1 infection. AIDS. 2001;15: 847–855. doi:10.1097/00002030-200105040-00005

48. Behrens G, Dejam A, Schmidt H, Balks HJ, Brabant G, Körner T, et al. Impaired glucose tolerance, beta cell function and lipid metabolism in HIV patients under treatment with protease inhibitors. AIDS. 1999;13: F63–70. doi:10.1097/00002030-199907090-00001

49. Cruz-Jentoft AJ, Bahat G, Bauer J, Boirie Y, Bruyère O, Cederholm T, et al. Sarcopenia: revised European consensus on definition and diagnosis. Age Ageing. 2019;48: 16–31. doi:10.1093/ageing/afy169

50. Chen L-K, Woo J, Assantachai P, Auyeung T-W, Chou M-Y, Iijima K, et al. Asian Working Group for Sarcopenia: 2019 Consensus Update on Sarcopenia Diagnosis and Treatment. J Am Med Dir Assoc. 2020;21: 300–307.e2. doi:10.1016/j.jamda.2019.12.012

51. da Silva Alexandre T, de Oliveira Duarte YA, Ferreira Santos JL, Wong R, Lebrão ML. Sarcopenia according to the european working group on sarcopenia in older people (EWGSOP) versus Dynapenia as a risk factor for disability in the elderly. J Nutr Health Aging. 2014;18: 547–553. doi:10.1007/s12603-014-0465-9

52. High KP, Brennan-Ing M, Clifford DB, Cohen MH, Currier J, Deeks SG, et al. HIV and Aging: State of Knowledge and Areas of Critical Need for Research. A Report to the NIH Office of AIDS Research by the HIV and Aging Working Group. JAIDS Journal of Acquired Immune Deficiency Syndromes. 2012;60: S1–S18. doi:10.1097/QAI.0b013e31825a3668

